# Suicidality and needs among people seeking medical assistance in dying based on psychological suffering: a protocol of a prospective longitudinal observational study

**DOI:** 10.1101/2025.05.08.25327143

**Authors:** Anoek Braam, Leah Middelkoop, Monique Kammeraat, Dik Meeuwis, Katja ten Cate, Sisco van Veen, Lizanne Schweren

## Abstract

**Introduction:** The number of requests for medical assistance in dying (MAID) based on psychological suffering has significantly increased in recent years. People that request MAID based on psychological suffering often have a long-term history within mental health care that has not (yet) led to recovery. The risk of death by suicide in this group is high. Relatively little is known about people who request MAID based on psychological suffering and their death wish. The lack of knowledge results in a lack of targeted counseling options and suicide prevention for people who request MAID based on psychological suffering. The current study therefore focuses on the characteristics, psychological processes and support needs of people who request MAID based on psychological suffering, and how death wishes and suicidality develop during and after a MAID request. The aim of this research is to contribute to better care and support for this vulnerable group.

**Methods:** To answer the questions above we conduct a prospective longitudinal observational study. Participants twice complete a digital questionnaire: one shortly after registering with Expertisecentrum Euthanasie (EE) for their MAID request and one six months after completing the first questionnaire. Participants are 16 years or older and have requested MAID at EE based on psychological suffering.

**Ethics and dissemination:** The study was approved as a study subject to the Medical Research Involving Human Subject Act (WMO) by the METC of Amsterdam UMC. Risks of participating in the study are low and explicit attention is paid to participant safety. The study results will be disseminated through scientific publication, conferences, and a webinar for mental health professionals. Based on participants support needs, a plan will be developed for support services to prevent suicide by people who requested MAID based on psychological suffering. Thus, the study will contribute to better counseling and suicide prevention for this vulnerable group.

## Introduction

Suicide prevention is an important public health priority.^1^ The risk of suicide is particularly high among vulnerable groups, such as people with severe, recurrent and/or persistent psychiatric disorders. In a limited number of European countries (Belgium, Switzerland, Luxembourg, Sweden, and the Netherlands), this group can seek medical assistance in dying (also referred to as euthanasia, assisted suicide or physician assisted death) based on psychological suffering (or: psychiatric MAID).^1^ In these countries, there has been a notable increase in both the number of requests and number of cases granted of assisted suicide among psychiatric patients.^1^ Despite the legal option for psychiatric MAID, research from Belgium and the Netherlands indicates that a minority of psychiatrists are willing to actively participate in the procedure.^2^ Psychiatrists find it challenging to explore a psychiatric MAID request as they consider it their professional role to provide hope and perspective to their patients.^3^

In the Netherlands, the increasing number of applications and the limited number of psychiatrists willing to be actively involved in the process has created an imbalance. As a result, the majority of psychiatric MAID requests is currently submitted to Expertisecentrum Euthanasie (Euthanasia Expertise Centre, the Netherlands [EE]).), the Dutch healthcare center providing MAID to patients who cannot get these services from their own doctors.^4^ To illustrate, in 2023 EE received 895 requests for psychiatric MAID,^4,5^ resulting in undesirable waiting times before processing of a request can be initiated. A recent study among young applicants (<24 years) with a psychiatric MAID request showed that the vast majority of them (>90%) either retracted their application or saw their application being rejected. Moreover, several patients died by suicide before processing of their application was completed, which can take months to over two years.^5^ It is important to note that suicide almost always results from a multitude of factors, including social isolation, challenging life circumstances and (untreated) mental health conditions. Prolonged waiting times may contribute by increasing feelings of hopelessness and exacerbating existing mental health conditions, although direct evidence of a relationship between suicide risk and waiting times is missing.^6,7^

The risk of suicide may also increase when a MAID-application is rejected. A Dutch study on patients requesting and receiving euthanasia between 2012 and 2018 indicates that 9.5% of psychiatric MAID requests were granted. In the same period, 19.6% of applicants withdrew their request and 42.6% of applicants received the message that their request could not be granted.^8^ Two studies show, despite considerable methodological limitations, that there is a higher suicide risk among people whose psychiatric MAID request is rejected. One study found that 16% of rejected applicants died by suicide, while another study reported a 12% suicide rate among rejected applicants.^9,10^

Taken together, people requesting psychiatric MAID are at greater risk of death by suicide, possibly moderated by increased hopelessness and despair, two key predictors of suicidal behavior.^11^ Furthermore, the reluctance of psychiatrists to discuss death as a potential outcome may reinforce patients’ feelings that their suffering is not recognized or taken seriously.^12^ Given these dynamics, it is crucial to develop and implement suicide prevention programs for this vulnerable group.

Unfortunately, effective suicide prevention among psychiatric MAID applicants is currently hindered by a gap in knowledge. First, it is crucial to gain understanding on the broad spectrum of death wishes as they occur among people with severe mental suffering. It is likely that many people with a death wish are also struggling with (phases of) acute suicidal thoughts and behaviors. Earlier research by Elzinga and colleagues highlighted that long-term death wishes can occur alongside (acute) suicidality, which may have a more variable, erratic, and impulsive character.^13^ Despite this being intuitively perceived by most participants in the study from Elzinga and colleagues, the distinction has not been systematically studied. Second, it is plausible that the help needs of the applicants for psychiatric MAID are different from others struggling with suicidal thoughts. For instance, applicants for psychiatric MAID often have extensive histories of (unsuccessful) mental health treatment trajectories. It is unclear how and to what extent death wishes and suicidality were addressed in those trajectories. As another example, contrary to many people struggling with suicidality, loved ones of people requesting psychiatric MAID are generally aware of the death wish of the applicant.^4^ It remains to be investigated whether this provides support and may contribute to suicide prevention. There remains a lack of understanding what help needs the group have and which help can be effective to prevent suicide.

This study aims to address the following questions:

- What clinical and demographic characteristics of applicants for psychiatric MAID are related to suicidality (thoughts, plans, suicidal attempts)?
- What cognitive, affective, social, and spiritual processes characterize death wishes, and suicidality in people requesting psychiatric MAID?
- Is there a need for help among people who request psychiatric MAID and what does this need for help look like?
- To what extent do death wishes and suicidality change in people requesting psychiatric MAID during and after a euthanasia request?

By investigating these questions, this study seeks to provide deeper understanding of the nature of death wishes and suicidality among psychiatric MAID applicants. Ultimately, this knowledge will contribute to designing a strategy to prevent suicide among people who request psychiatric MAID and support these individuals more effectively. The group of people requesting psychiatric MAID is growing and an international subject of intense discussion. The availability of legal MAID for people struggling with psychiatric disorders in the Netherlands provides the opportunity to conduct a study among these people.

## Methods and analysis

From June 2024 to December 2025 we will conduct a prospective longitudinal observational study to provide deeper understanding of the nature of death wishes and suicidality among psychiatric MAID applicants at EE. Participants will twice receive a digital questionnaire: an initial measurement within one week after registering at EE and a repeated measurement 180 days after participating in the initial measurement. It is expected that after 180 days, most people who have registered with EE will have passed the triage at the medical records office.

This research is conducted by 113 Suicide Prevention, which is the Dutch organization for suicide prevention, as sponsor and main applicant, and EE as co-applicant. The study is subsidized by Stichting tot Steun VCVGZ. This foundation funds research and innovative projects that can contribute to mental health care.

### Study population

Recruitment of participants will take place at the Dutch MAID expertise center (EE). EE is not a physical place where people can go for MAID. Instead EE acts as an intermediary for people who cannot turn to their own practitioner for MAID. In recent years, an estimated average of 55% of all psychiatric MAID procedures were performed by EE.^14–16^ The study population includes all people aged 16 years and older who apply to EE to be considered for psychiatric MAID, and who have consented to be approached for scientific research. Based on prior years, EE is expected to receive 800-1000 applications for psychiatric MAID within the study period.

### Care as usual

Psychiatric MAID applications follow a multi-step process to determine a patient’s eligibility for MAID. At each step, patients either discontinue their applications (due to various reasons, including death by suicide or natural causes), are rejected, or move on to the next step. While administrative procedures have evolved over time to accommodate growing demand, the overall process remains consistent. The application steps can generally be described as follows.

The initial request consists of the patient (or, at the patients request, their doctor or relative) submitting an application form that outlines the patient’s medical history and reasons for seeking MAID. Following this, the patient and their healthcare provider are required to submit comprehensive medical records, resulting in a complete application. The next step involves an in-person eligibility screening by a psychiatrist. If the application appears to meet the eligibility criteria, it proceeds to the full assessment phase, which often involves a waiting period. During this phase, to confirm the irremediability of the patients suffering, additional treatments may be required. The full assessment, which can take months to several years to complete, ultimately results in an eligibility decision, and if approved, the provision of MAID.

### Sample size calculation

For calculating the sample size, the following assumptions were used: 80% of applicants will consent to being approached for scientific research, from these 50% will respond to the first questionnaire, and from these respondents 50% will respond to the second questionnaire. This results in an expected sample size of N=320-400 respondents for the first measurement and N=160-200 respondents for the second measurement.

### Questionnaire

The questionnaires for measurement at time-point one and time-point two consists of items about the psychiatric history, the psychiatric MAID request, suicidality, characteristics that may be associated with a (persistent) death wish, and the desired or unmet help needs. The questionnaires combine six validated scales (51 items) with a number of items (27 items, of which 14 are conditional) designed for the current study. The demographic data and psychiatric history is only collected at the first time-point (10 items, of which 2 are conditional). Completion of each questionnaire takes an estimated 30 minutes.

The validated scales used are: (i) the Suicidal Subscale of the Depressive Symptom Index to assess thought of deaths,^17^ (ii) the Fearlessness about Death-Scale (GCSQ-FAD) of the German Capability for Suicide Questionnaire to assess fear about death,^18^ (iii) the Interpersonal Needs Questionnaire to assess the connectedness with other people,^19^ (iv) the Defeat and Entrapment Scale – to assess the feelings or thought of being stuck and defeated,^20^ (v) the short version of the Beck Hopelessness Scale to assess feelings of hope,^21^ and (vi) the Positive and Negative Framework subscale from the Life Regard Index to assess feelings of meaning.^22^

Other items are both open-ended and multiple choice items that concern the self-perceived origin(s) of the death wish, past and current psychiatric diagnoses, past and current treatment trajectories, history of suicidal behaviors and unmet help needs. Demographic items address age, gender and education.

### Data analysis plan

The first part of the study is descriptive in nature, namely the mapping of death wishes and suicidality among people applying for psychiatric MAID. We describe numbers and percentages for all categorical variables, and averages and distributions of all continuous variables. Help needs related to death wishes and suicidality are also described in this way.

The second part of the study is comparative in nature. We will compare groups of applicants based on the status of their application at the time of the second measurement (withdrawn, rejected, waiting list). The above descriptive analyses are repeated for each of the three groups. Repeated measures ANOVAs are performed, comparing changes between the two measurement times within the three groups. We will report both between-group and within-group effects.

The third part of the study is data-driven in nature. Provided that we achieve to include a sufficient number of patients in the study, a latent profile analysis (LPA) will be performed on the sum scores related to all characteristics at the time of application, with the aim of identifying clinically relevant subgroups among people requesting psychiatric MAID. For the LPA, all relevant model parameters will be reported. Descriptive analysis of all identified subgroups will be presented regarding demographic and clinical characteristics, and case status at the time of the second measurement. These characteristics will be additionally compared between the identified groups, using ANOVAs for continuous variables and Chi^2^ test for categorical variables.

Regarding the open-ended questions, these results are qualitative in nature and will be analyzed through inductive thematic coding.

### Ethics

The study has been approved by the Medical Ethical Review Committee of the Amsterdam University Medical Center (METC 2023.1008) and will be conducted in accordance with relevant laws and regulations and the principles of the Declaration of Helsinki.^23^ All patients will provide informed consent via the Castor eConsent environment – Castor is online software for digitizing consent forms and questionnaires.

#### Participant safety

The benefits of participation are negligible, though participants may feel relieved by sharing their experiences. Participants receive gift cards worth 10 euro’s after completing the first questionnaire and 15 euro’s after completing the second questionnaire. The risks of participation are expected to be low as several international and national studies show that questioning people about suicidality has no harmful effects.^24,25^ Despite a low risk assessment those who request psychiatric MAID have an increased risk of dying by suicide. To minimize the risk of dying by suicide the general practitioner or mental health practitioner of the participant is informed about their patient’s participation in the study. Furthermore, participants are at the start and at the end of each questionnaire made aware of the possibility of free and anonymous contact with helpers at 113 Suicide Prevention, and aftercare is offered the moment a participant indicates that he or she experiences negative effects as a result of participating in the study.

#### Dissemination

Results of the study will be disseminated through several outlets, including reports to the accredited METC and funder of the study, peer-reviewed publications, and presentations at (inter)national conferences by Expertisecentrum Euthanasie and 113 Suicide Prevention and. Furthermore, the findings will be disseminated via meta-organizations within Dutch mental health care (Supranet GGZ and ThaNet).

Furthermore, the research will result in a plan for the development of support services aimed at preventing suicide among those applying for psychiatric MAID. The plan will outline the types of support that are needed and how these can be addressed.

## Conclusion

This paper presents a prospective longitudinal observational study that delves into the complex nature of death wishes, suicidality, and support needs among psychiatric MAID applicants. By tracking participants over time, the study seeks to uncover patterns and underlying factors that contribute to the emergence and persistence of such thoughts within this vulnerable population. The ultimate aim of the research is to develop targeted strategies to support psychiatric MAID applicants, and thereby reduce the risk of suicide in this group.

Findings from the study will be critical for the evidence-based development of support services that meet the needs of psychiatric MAID applicants. These insights will also equip healthcare professionals, particularly psychiatrists, with necessary knowledge and tools to provide care to individuals that apply for psychiatric MAID and foster a more nuanced understanding of patient experiences and needs.

Moreover, this research fills a significant gap in literature by offering empirical data that can guide future interventions in the realm of psychiatric care, suicide prevention, and end-of-life choices. It holds the potential to not only mitigate the risk of suicide but also to enhance the holistic support offered to patients, ensuring that their physical, psychological, and existential concerns are addressed with empathy.

## Data Availability

All data produced based on this study protocol will be available upon reasonable request to the authors

## Funding statement

This work was supported by Stichting tot Steun Vereniging voor Christelijke Verzorging van Geestes-en Zenuwzieken (Stichting tot Steun VCVGZ).

## Competing interests statement

None declared.

## Notes

### Competing Interest Statement

The authors have declared no competing interest.

### Author Declarations

The Medical Ethical Review Committee of the Amsterdam University Medical Center gave ethical approval for this work

### Summary of Updates

Introduction. The number of requests for medical assistance in dying (MAID) based on psychological suffering has significantly increased in recent years. People that request MAID based on psychological suffering often have a long-term history within mental health care that has not (yet) led to recovery. The risk of death by suicide in this group is high. Relatively little is known about people who request MAID based on psychological suffering and their death wish. The lack of knowledge results in a lack of targeted counseling options and suicide prevention for people who request MAID based on psychological suffering. The current study therefore focuses on the characteristics, psychological processes and support needs of people who request MAID based on psychological suffering, and how death wishes and suicidality develop during and after a MAID request. The aim of this research is to contribute to better care and support for this vulnerable group. Methods. To answer the questions above we conduct a prospective longitudinal observational study. Participants twice complete a digital questionnaire: one shortly after registering with Expertisecentrum Euthanasie (EE) for their MAID request and one six months after completing the first questionnaire. Participants are 16 years or older and have requested MAID at EE based on psychological suffering. Ethics and dissemination. The study was approved as a study subject to the Medical Research Involving Human Subject Act (WMO) by the METC of Amsterdam UMC. Risks of participating in the study are low and explicit attention is paid to participant safety. The study results will be disseminated through scientific publication, conferences, and a webinar for mental health professionals. Based on participants support needs, a plan will be developed for support services to prevent suicide by people who requested MAID based on psychological suffering. Thus, the study will contribute to better counseling and suicide prevention for this vulnerable group.

## References

1. Calati R, Olié E, Dassa D, et al. Euthanasia and assisted suicide in psychiatric patients: A systematic review of the literature. J Psychiatr Res. 2021;135:153–173. doi:10.1016/j.jpsychires.2020.12.006

2. Verhofstadt M, Audenaert K, Van Den Broeck K, et al. Belgian psychiatrists’ attitudes towards, and readiness to engage in, euthanasia assessment procedures with adults with psychiatric conditions: a survey. BMC Psychiatry. 2020;20(1):374. doi:10.1186/s12888-020-02775-x

3. Besjes MJ, van de Vathorst S. Euthanasie in de ggz: kwalitatief onder-zoek naar de mening van psychiaters. Tijdschr VOOR Psychiatr. Published online 2023.

4. Kammeraat M, Van Rooijen G, Kuijper L, Kiverstein JD, Denys DAJP. Patients requesting and receiving euthanasia for psychiatric disorders in the Netherlands. BMJ Ment Health. 2023;26(1):e300729. doi:10.1136/bmjment-2023-300729

5. Schweren LJS, Rasing SPA, Kammeraat M, et al. Requests for Medical Assistance in Dying by Young Dutch People With Psychiatric Disorders. JAMA Psychiatry. 2025;82(3):246–252. doi:10.1001/jamapsychiatry.2024.4006

6. Lange wachttijden in de GGZ: wat doen we in de tussentijd? De EO. Accessed October 14, 2024. https://www.eo.nl/artikel/lange-wachttijden-in-de-ggz-wat-doen-we-in-de-tussentijd

7. CDC. Risk and Protective Factors for Suicide. Suicide Prevention. May 20, 2024. Accessed October 14, 2024. https://www.cdc.gov/suicide/risk-factors/index.html

8. Onderzoeksrapportage-Psychiatrische-Patiënten-Expertisecentrum-Euthanasie.pdf. Accessed October 21, 2024. https://expertisecentrumeuthanasie.nl/app/uploads/2020/02/Onderzoeksrapportage-Psychiatrische-Pati%C3%ABnten-Expertisecentrum-Euthanasie.pdf

9. Derde_evaluatie_Wtl.pdf. Accessed June 11, 2024. https://www.zonmw.nl/sites/zonmw/files/typo3-migrated-files/Derde_evaluatie_Wtl.pdf

10. Thienpont L, Verhofstadt M, Van Loon T, Distelmans W, Audenaert K, De Deyn PP. Euthanasia requests, procedures and outcomes for 100 Belgian patients suffering from psychiatric disorders: a retrospective, descriptive study. BMJ Open. 2015;5(7):e007454. doi:10.1136/bmjopen-2014-007454

11. Risicofactoren - 4. Suïcidaliteit - Richtlijn Stemmingsproblemen. Richtlijnen jeugdhulp en jeugdbescherming. Accessed September 17, 2024. https://richtlijnenjeugdhulp.nl/stemmingsproblemen/suicidaliteit/risico-instandhoudende-en-beschermende-factoren/

12. Pronk R, Willems DL, van de Vathorst S. Feeling Seen, Being Heard: Perspectives of Patients Suffering from Mental Illness on the Possibility of Physician-Assisted Death in the Netherlands. Cult Med Psychiatry. 2022;46(2):475–489. doi:10.1007/s11013-021-09726-5

13. Elzinga E, Veen S van, Wijngaarden E van, et al. Een levenslange doodswens en persisterende suïcidaliteit: een exploratief onderzoek naar het ontstaan, verloop en behoeften van mensen die hieraan lijden. Published online 2021. Accessed June 11, 2024. https://researchinformation.amsterdamumc.org/en/publications/een-levenslange-doodswens-en-persisterende-su%C3%AFcidaliteit-een-expl

14. Regionale Toetsingscomissies Euthanasie Jaarverslag 2022. file:///C:/Users/AnoekBraam/Downloads/RTE_JV2022_def_2april.pdf

15. Regionale Toetsingscommissies Euthanasie Jaarverslag 2023. file:///C:/Users/AnoekBraam/Downloads/RTE-Jaarverslag-2023-defst.pdf

16. Regionale Toetsingscommissies Euthanasie Jaarverslag 2024. file:///C:/Users/AnoekBraam/Downloads/RTE-jaarverslag2024-DEF+17-3-25.pdf

17. Psychometric properties of the Depressive Symptom Index-Suicidality Subscale (DSI-SS) in an adult psychiatric sample. Accessed July 17, 2024. https://psycnet.apa.org/doiLanding?doi=10.1037%2Fpas0001043

18. Validation of the German capability for suicide questionnaire (GCSQ) in a high-risk sample of suicidal inpatients | BMC Psychiatry | Full Text. Accessed July 17, 2024. https://bmcpsychiatry.biomedcentral.com/articles/10.1186/s12888-020-02812-9

19. Marty MA, Segal DL, Coolidge FL, Klebe KJ. Analysis of the psychometric properties of the Interpersonal Needs Questionnaire (INQ) among community-dwelling older adults. J Clin Psychol. 2012;68(9):1008–1018. doi:10.1002/jclp.21877

20. Griffiths AW, Wood AM, Maltby J, Taylor PJ, Panagioti M, Tai S. The development of the Short Defeat and Entrapment Scale (SDES). Psychol Assess. 2015;27(4):1182–1194. doi:10.1037/pas0000110

21. Kliem S, Lohmann A, Mößle T, Brähler E. Psychometric properties and measurement invariance of the Beck hopelessness scale (BHS): results from a German representative population sample. BMC Psychiatry. 2018;18(1):110. doi:10.1186/s12888-018-1646-6

22. Debats DLHM. Meaning in Life: Psychometric, Clinical and Phenomenological Aspects. s.n.; 1996.

23. World Medical Association. World Medical Association Declaration of Helsinki: Ethical Principles for Medical Research Involving Human Subjects. JAMA. 2013;310(20):2191–2194. doi:10.1001/jama.2013.281053

24. DeCou CR, Schumann ME. On the Iatrogenic Risk of Assessing Suicidality: A Meta-Analysis. Suicide Life Threat Behav. 2018;48(5):531–543. doi:10.1111/sltb.12368

25. Bender TW, Fitzpatrick S, Hartmann MA, et al. Does it hurt to ask? An analysis of iatrogenic risk during suicide risk assessment. Neurol Psychiatry Brain Res. 2019;33:73–81. doi:10.1016/j.npbr.2019.07.005

